# COVID-19 severity and risk of SARS-CoV-2-associated asthma exacerbation by time since booster vaccination: a longitudinal analysis of data from the COVIDENCE UK study

**DOI:** 10.1101/2024.06.28.24309666

**Authors:** Giulia Vivaldi, Mohammad Talaei, Paul E Pfeffer, Seif O Shaheen, Adrian R Martineau

## Abstract

**Background:** In several countries, COVID-19 booster vaccinations are offered annually to priority groups, but many people have not been vaccinated in over a year. We aimed to assess the association between time since booster vaccination and characteristics of breakthrough infection. We also assessed whether incident COVID-19 continued to associate with asthma exacerbations in boosted individuals, and whether risk of COVID-19-associated exacerbation was affected by time since vaccination.

**Methods:** COVIDENCE UK is a prospective, longitudinal, population-based study of COVID-19. We included adult participants who had received ≥1 booster vaccination. Time since vaccination was binarised at 6 months or 12 months according to vaccine eligibility subgroup. We used logistic, Cox, and linear regression to obtain adjusted estimates for the association between time since vaccination and breakthrough infection severity, symptom duration, and acute changes to health-related quality of life (measured by the EQ-5D-3L Index). We then assessed the association of incident COVID-19 with asthma exacerbations using multilevel mixed models, by time since vaccination.

**Results:** 7391 boosted participants reported a breakthrough infection. Across all eligibility subgroups, greater time since vaccination associated with increased odds of infection requiring bedrest (*vs* milder symptoms), with the highest odds for adults aged 65–75 years (1.83 [95% CI 1.51–2.23] when vaccinated >6 months *vs* ≤6 months prior). However, we observed little evidence of association between time since vaccination and symptom duration. Vaccination >12 months prior (*vs* ≤12 months) was associated with a small decrease in EQ-5D-3L Index among participants younger than 65 years (-0.03 points [-0.04 to -0.01]). Among 2100 participants with asthma, incident COVID-19 associated with increased risk of asthma exacerbation, both ≤12 months after vaccination (OR 5.31 [4.36–6.48]) and later (6.06 [3.23–11.38]), with a greater difference in point estimates when specifically considering severe asthma exacerbations (6.82 [4.88–9.54] for ≤12 months *vs* 10.06 [3.90–25.92] for >12 months).

**Conclusion:** Longer time since booster vaccination consistently associates with more severe breakthrough infections, and may potentially increase risk of severe asthma exacerbations. These findings highlight the importance of ensuring those currently eligible receive their booster vaccinations, and the need for research on further vaccinations in people with asthma no longer eligible for boosters.

## Introduction

More than 3 years since the start of vaccination programmes against SARS-CoV-2, booster vaccinations continue to be offered in countries across the world. [1] Within the context of Omicron dominance and high population immunity, the WHO has published recommendations of priority groups for repeated booster vaccinations; [2] these recommendations are broadly followed by several countries, [3-5] with others continuing to offer annual vaccinations for everyone. [6]

In the UK, booster vaccinations are offered annually to priority groups in the autumn, [7] with more than 7 million people aged 65 years and older vaccinated in autumn 2023. [8] Some more vulnerable groups, such as adults aged 75 years and older, care home residents, and the immunosuppressed, are additionally offered a vaccine in the spring. [7] However, eligibility for SARS-CoV-2 booster vaccinations is continually under review, [5] with the offering reduced each autumn to date. [7]

Vaccine effectiveness is known to wane over time; [9, 10] therefore, longer time since vaccination, either because of vaccine ineligibility or under-vaccination, [11] may put people at increased risk of infection and severe disease. Vaccine protection against severe outcomes lasts longer than protection against mild disease; [12] however, mild-to-moderate COVID-19 remains an important concern, with acute COVID-19 and long-term sequelae continuing to be a major reason for sickness absence in the workforce. [13] In people with breakthrough SARS-CoV-2 infection, greater time since vaccination has been shown to associate with greater symptom severity [14, 15] and increased risk of severe or critical COVID-19. [16-19] However, few studies have looked at symptom duration [14], and no studies, to our knowledge, have assessed how time since booster vaccination may affect the infection’s impact on health-related quality of life. Additionally, very few studies have considered outcomes more than 8 months after booster vaccinations, [16, 20] whereas an increasing portion of the UK population has not received a SARS-CoV-2 vaccine for a year or more. [7]

Among those currently eligible for annual booster vaccination are people with poorly controlled asthma. [7] In this population, the protective effect of vaccination is twofold: on the one hand, vaccination protects against severe COVID-19, for which poorly controlled asthma is a risk factor; [21] on the other, by reducing the incidence and severity of breakthrough infections, vaccination may also protect against asthma exacerbations triggered by incident COVID-19 [22, 23] and subsequent worsening of asthma control. [24] However, existing studies into the relationship between incident COVID-19 and asthma exacerbations have not been carried out in boosted populations. [22, 23, 25] Additionally, these studies have not considered whether exacerbation risk from breakthrough SARS-CoV-2 infection is affected by time since vaccination, which is likely to be longer in people with well controlled asthma, who may still experience severe asthma exacerbations [26] but are currently not eligible for booster vaccinations. [7]

To address these knowledge gaps, we therefore aimed to assess the association between time since vaccination and various characteristics of breakthrough infections, including symptom severity, symptom duration, and acute changes to health-related quality of life. We also aimed to evaluate whether incident COVID-19 continued to be associated with asthma exacerbations in a boosted population, and whether this association was affected by time since booster vaccination.

## Methods

### Study design and participants

COVIDENCE UK is a prospective, longitudinal, population-based observational study of COVID-19 in the UK population (https://www.qmul.ac.uk/covidence).^15^ Inclusion criteria were age 16 years or older and UK residence at enrolment, with no exclusion criteria. Participants were invited via a national media campaign via print and online newspapers, radio, television, social media, and online advertising. Enrolled participants completed an online baseline questionnaire and monthly follow-up questionnaires to capture information on potential symptoms of COVID-19, results of nose or throat swab tests for SARS-CoV-2, vaccinations, and asthma exacerbations. The study was launched on May 1, 2020, and closed to enrolment on Oct 6, 2021. The final COVIDENCE UK cohort was majority female (70.2%) and White (93.7%), with under-representation of people younger than 50 years, men, and minority ethnicities.^15^ This study is registered with ClinicalTrials.gov, NCT04330599. COVIDENCE UK was approved by Leicester South Research Ethics Committee (ref 20/EM/0117). All participants gave informed consent to take part in the study before enrolment.

In this analysis, we included all participants who had received at least one COVID-19 booster vaccination and reported a positive nose or throat swab test for SARS-CoV-2 at least 14 days after their last vaccination. This analysis was pragmatic in nature, including all participants meeting these eligibility criteria with no sample size specified.

### SARS-CoV-2 infection

We defined evidence of a SARS-CoV-2 infection as a positive SARS-CoV-2 swab test. Where participants reported more than one post-booster SARS-CoV-2 infection, we focused on the last infection reported, to increase representation of longer times since last vaccination. Infection dates were defined as the date of the positive test. Infections reported in the 14 days following a vaccination were excluded. Participants with symptoms were asked to report the severity of their symptoms as mild (able to do most of usual activities), moderate (unable to do usual activities, but without requiring bedrest), or feeling very unwell (requiring bedrest). Participants were asked to report their symptom onset date and whether they felt back to normal on the day of the questionnaire; if they responded yes, they were asked to report a symptom end date. Participants not reporting symptoms were classified as asymptomatic.

### Booster vaccine eligibility groups

After the emergence of Omicron, all adults in the UK were eligible for the first round of booster vaccinations, which began in autumn 2021; however, additional booster vaccinations have been restricted to specific subgroups according to their risk of severe outcomes, their contact with potentially vulnerable people, and to protect NHS capacity. [7] The requirement for further vaccinations is regularly reviewed by the Joint Committee on Vaccination and Immunisation, [7] leading to some groups no longer being eligible for further vaccinations (Figure 1).

**Figure 1:**
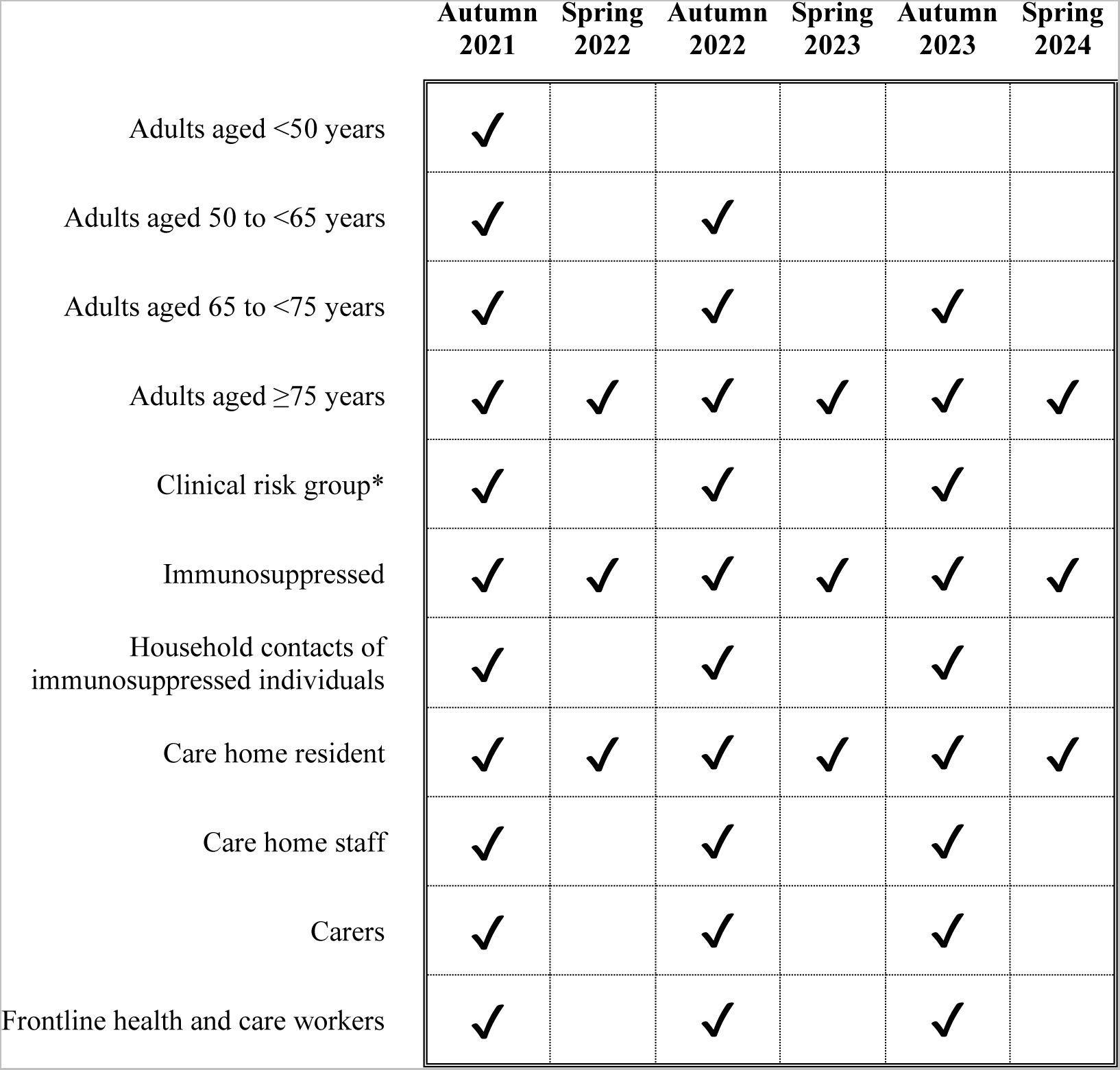
Vaccine eligibility. *The clinical risk group defined by the Joint Committee on Vaccination and Immunisation is comprised of individuals with a wide range of health conditions, outlined in Table S 1.

COVIDENCE UK does not have data on all of the eligibility criteria for the clinical risk group; we therefore classified participants as being in the clinical risk group if they reported chronic obstructive pulmonary disease or poorly controlled asthma (based on medications taken or exacerbations reported in monthly questionnaires; see Table S 1), diabetes, heart disease, peripheral vascular disease, kidney disease, major neurological conditions, immunosuppression, morbid obesity, or splenectomy.

### Asthma exacerbations

Participants were asked to report asthma exacerbations occurring in the past month. Exacerbations were classified as severe if they required treatment with systemic corticosteroids or precipitated emergency department attendance or hospital admission.

### Time since booster vaccination

Time since booster vaccination was defined as the number of days between the last vaccine received and the infection date. To assess whether more frequent vaccination would be beneficial for particular subgroups, we converted time since vaccination into a binary variable, classifying participants as having received a vaccine in the past 6 months (for those currently eligible for annual vaccination, but not 6-monthly vaccination) or the past 12 months (for those currently not eligible for annual vaccination).

### Covariates

Potential confounders of the association between time since vaccination and breakthrough COVID-19 severity were identified using a directed acyclic graph (Figure 2). While many risk factors have been identified for greater COVID-19 severity after vaccination, [27, 28] we specifically considered those that might affect vaccine eligibility and vaccine uptake, which could both impact time since last vaccination. Owing to the aims of the UK vaccine programme, most of the factors that affect vaccine eligibility also affect disease severity. [7] Vaccine uptake or hesitancy is associated with various factors not captured in COVIDENCE UK, such as compliance with health restrictions and engaging with conspiracy theories; [29] however, it has also been found to associate with factors such as age, sex, general health, and socioeconomic status. [30] We therefore adjusted for age and other vaccine eligibility criteria (ie, comorbidities and frontline worker status), sex, ethnicity, socioeconomic status (represented by level of educational attainment and Index of Multiple Deprivation), general health, previous SARS-CoV-2 infection, and the number of vaccines received at time of infection.

**Figure 2:**
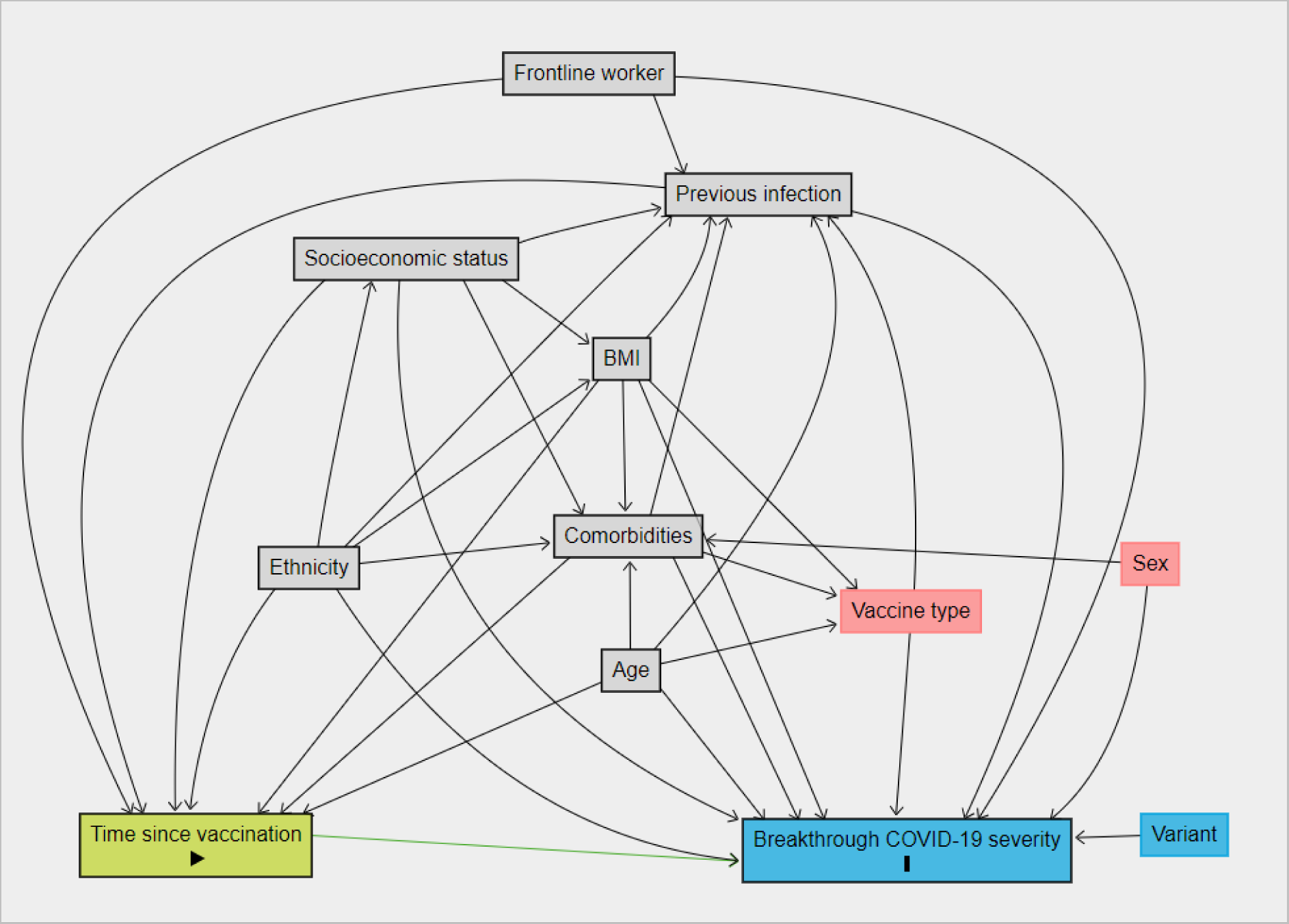
Directed acyclic graph for the association between time since vaccination and breakthrough COVID-19 severity

### Statistical analysis

First, we assessed the potential impact of time since vaccination on three dimensions of SARS-CoV-2 infection: reported symptom severity, duration of symptoms, and health-related quality of life.

We used logistic regression to explore the association between self-reported symptom severity and time since vaccination, converting severity into a binary variable (very unwell *vs* all other severities). We compared differences in symptom duration using Cox regression, censoring participants at the questionnaire date if they did not report a symptom end date for that infection episode. Finally, we examined changes in health-related quality of life using the EQ-5D-3L, [31] which participants complete in each monthly questionnaire. We converted the five dimensions of the EQ-5D-3L into a weighted health state index, using EQ-5D preference weights obtained from the UK general population, [32] and calculated the difference between the average value in the 6 months before infection and the value in the month immediately following the infection. We then assessed the association between the change in EQ-5D-3L index and time since vaccination using linear regression.

Second, we evaluated whether incident SARS-CoV-2 infection continued to be associated with asthma exacerbations in a boosted population, and whether this association was affected by time since vaccination. We included all boosted participants with asthma and post-booster follow-up. We excluded any observations in the first month after a SARS-CoV-2 vaccination as we could not guarantee that any exacerbations reported would have occurred after the vaccination. We calculated odds ratios (ORs) for associations between incident SARS-CoV-2 infection and both mild and severe asthma exacerbations using multilevel logistic generalised linear models, with random effects for participants. We included SARS-CoV-2 infection as a three-level independent variable: no infection, infection when vaccinated in the previous 12 months, and infection when vaccinated more than 12 months prior. We adjusted for the same covariates as in the SARS-CoV-2 infection models, while additionally adjusting for asthma treatment level (use of reliever inhaler only, use of inhaled corticosteroid, use of a long-acting bronchodilator inhaler, or monoclonal antibody therapy), asthma exacerbation history, incident non-COVID-19 acute respiratory infection, tobacco smoking status, season (October–April *vs* May–September), and measures of response rate and SARS-CoV-2 testing rate.

We did two sensitivity analyses. First, we restricted the analyses to participants with previous infection in order to control for the severity of their most severe previous infection. In doing so, we aimed to factor in participants’ predisposition towards severe infections, which may not be fully accounted for by the covariates available to us. Second, as the analyses are restricted to people who report positive tests results, we are effectively conditioning on the likelihood of being infected and taking a SARS-CoV-2 test, putting us at risk of collider bias. [33] We therefore carried out inverse-probability weighted regressions with robust standard errors, with weights based on the probability of reporting a positive test calculated on all participants who had received a booster vaccination.

The proportional hazards assumption for the Cox models was tested using Schoenfeld residuals and visual assessment of log–log plots of survival; models were then stratified by variables found to violate the assumption to assess the impact on our estimates. Missing data were handled with listwise deletion. All analyses were done with Stata 18.0.

## Results

Between August, 2021, and January, 2024, 7391 boosted participants reported a SARS-CoV-2 infection (Figure 3). Participants had a median age of 63 years (IQR 55–69), 5368 (72.6%) were female, and 7139 (96.6%) were White (Table 1). 2369 (32.1%) participants reported being very unwell during their post-booster infection: 1254 (28.0%) of 4483 participants vaccinated in the previous 6 months, 915 (37.6%) of 2432 vaccinated 6–12 months prior, and 200 (42.0%) of 476 vaccinated more than 1 year prior. The number of vaccines received before post-booster infection was similar between severity groups (Table 1). In this boosted population, more than 99% of infections occurred after the Omicron variant became dominant (Dec 17, 2021) [7] and less than 0.5% of infections led to hospitalisation.

**Figure 3:**
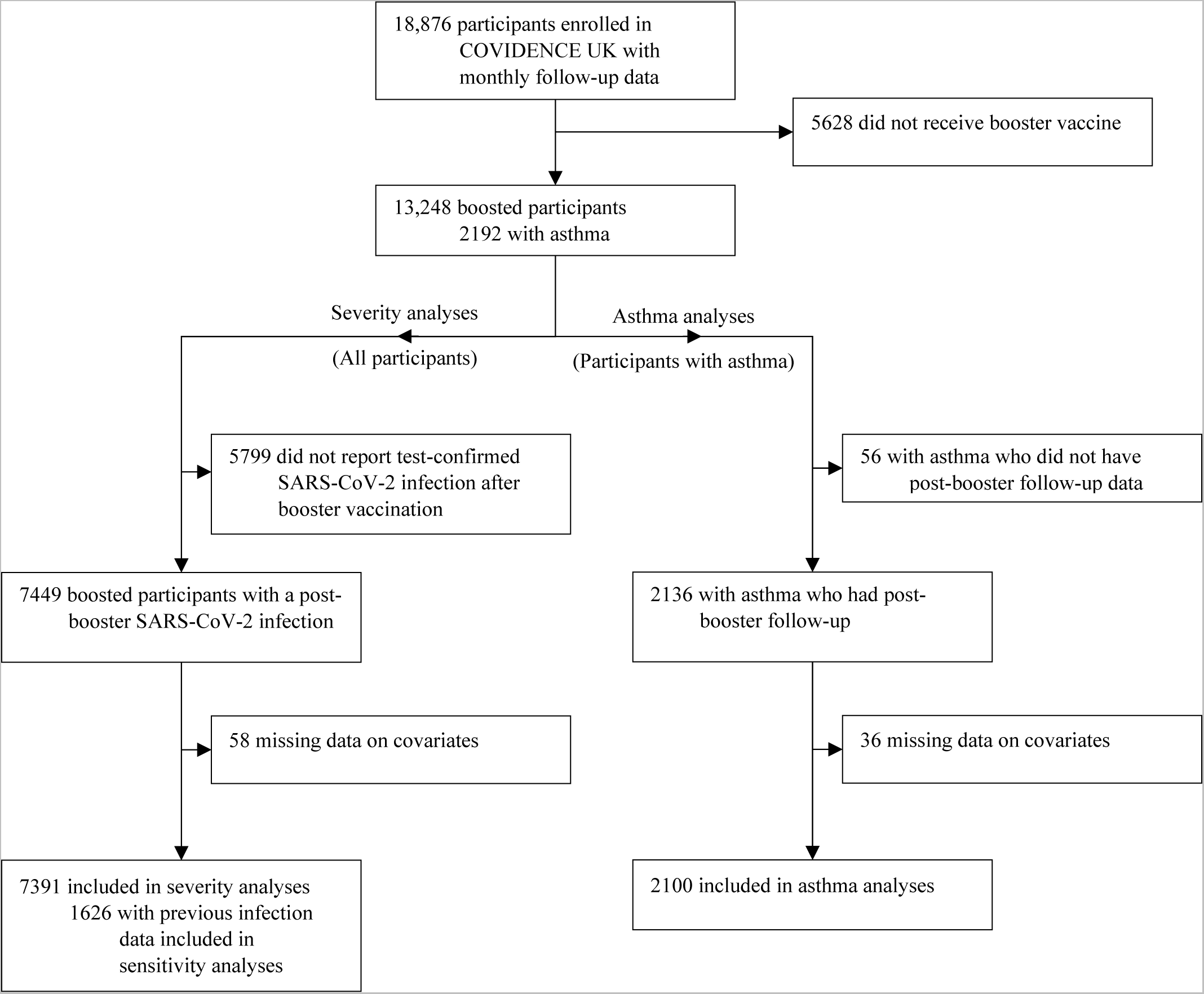
Participant flow diagram

**Table 1:** Participant characteristics, by severity of post-booster COVID-19.

|  | Very unwell<br>(N=2369) | Asymptomatic, mild, or<br>moderate (N=5022) |
| --- | --- | --- |
| **Sociodemographics** |  |  |
| Age, years | 60 (51–67) | 64 (57–70) |
| <50 | 498 (21.0%) | 695 (13.8%) |
| 50 to <65 | 1088 (45.9%) | 2057 (41.0%) |
| 65 to <75 | 665 (28.1%) | 1882 (37.5%) |
| ≥75 | 118 (5.0%) | 388 (7.7%) |
| Sex |  |  |
| Female | 1895 (80.0%) | 3473 (69.2%) |
| Male | 474 (20.0%) | 1549 (30.8%) |
| Ethnicity |  |  |
| Black, African, Caribbean, or Black British | 3 (0.1%) | 23 (0.5%) |
| South Asian | 30 (1.3%) | 49 (1.0%) |
| White | 2273 (95.9%) | 4866 (96.9%) |
| Mixed, multiple, or other ethnic groups | 63 (2.7%) | 84 (1.7%) |
| Quartiles of IMD decile |  |  |
| Q4 (least deprived) | 788 (33.3%) | 1778 (35.4%) |
| Q3 | 637 (26.9%) | 1331 (26.5%) |
| Q2 | 475 (20.1%) | 1001 (19.9%) |
| Q1 (most deprived) | 469 (19.8%) | 912 (18.2%) |
| Highest educational level attained |  |  |
| Primary or secondary | 186 (7.9%) | 492 (9.8%) |
| Higher or further (A levels) | 295 (12.5%) | 696 (13.9%) |
| College or university | 1044 (44.1%) | 2286 (45.5%) |
| Post-graduate | 844 (35.6%) | 1548 (30.8%) |
| Frontline health or care worker | 242 (10.2%) | 423 (8.4%) |
| Booster vaccines received |  |  |
| 1 | 1546 (65.3%) | 3252 (64.8%) |
| 2 | 600 (25.3%) | 1267 (25.2%) |
| ≥3 | 223 (9.4%) | 502 (10.0%) |
| **Infection details** |  |  |
| Symptom duration, days | 22 (13–33) | 12 (7–23) |
| Time since last vaccination, days | 188 (112–280) | 153 (91–241) |
| Vaccinated in past 6 months | 1254 (52.9%) | 3229 (64.3%) |
| Vaccinated in past 12 months | 2169 (91.6%) | 4746 (94.5%) |
| Any previous infections | 478 (20.2%) | 1155 (23%) |
| Previous infection severity |  |  |
| Asymptomatic | 15/474 (3.2%) | 77/1142 (6.7%) |
| Mildly unwell | 70/474 (14.8%) | 367/1142 (32.1%) |
| Moderately unwell | 123/474 (25.9%) | 408/1142 (35.7%) |
| Very unwell | 266/474 (56.1%) | 290/1142 (25.4%) |

*Table 1 continues on next page*
|  | **Very unwell**<br>**(N=2369)** | **Asymptomatic, mild, or**<br>**moderate (N=5022)** |
| --- | --- | --- |
| **Clinical characteristics** |  |  |
| BMI, kg/m<sup>2</sup> |  |  |
| <25 | 1131 (47.7%) | 2600 (51.8%) |
| 25 to <30 | 729 (30.8%) | 1608 (32.0%) |
| ≥30 | 509 (21.5%) | 814 (16.2%) |
| Asthma | 517 (21.8%) | 754 (15.0%) |
| Diabetes | 99 (4.2%) | 210 (4.2%) |
| Heart disease | 92 (3.9%) | 189 (3.8%) |
| Kidney disease | 50 (2.1%) | 94 (1.9%) |
| Hypertension | 466 (19.7%) | 1141 (22.7%) |
| Systemic immunosuppressants | 156 (6.6%) | 241 (4.8%) |
| Oral corticosteroids | 97 (4.1%) | 123 (2.4%) |
| Clinical risk group | 679 (28.7%) | 1206 (24.0%) |
| COPD or poorly controlled asthma | 339 (14.3%) | 480 (9.6%) |

Data are n (%), n/N (%), or median (IQR). Very unwell was defined as requiring bedrest. BMI=body-mass index. COPD=chronic obstructive pulmonary disease. IMD=Index of Multiple Deprivation.

1330 (18.0%) participants were eligible for 6-monthly vaccination and 5392 (73.0%) were initially eligible for annual vaccination; after the change in eligibility criteria for the autumn 2023 campaign, this number reduced to 3819 (51.7%), leaving 2242 (30.3%) participants ineligible for any further booster vaccinations. Approximately 65% of participants reported receiving all booster vaccinations they were eligible for.

Across all eligibility groups, greater time since vaccination was associated with increased odds of reporting being very unwell, although the small number of participants with controlled asthma compromised the statistical power of the analysis (Table 2). Although point estimates consistently suggested that greater time since vaccination might be associated with a longer time to recovery for some groups, evidence was weak (Table 2). Less recent vaccination was associated with a small decrease in health-related quality of life among participants younger than 65 years (Table 2).

**Table 2:** Associations between COVID-19 severity, speed of recovery, and health-related quality of life vs time since vaccination.

|  | Severity: very unwell* |  | Speed of recovery |  | Change in EQ-5D index |  |
| --- | --- | --- | --- | --- | --- | --- |
|  | OR (95% CI) | p value | HR (95% CI) | p value | Beta (95% CI) | p value |
| **Association with not having received a booster vaccination in previous 12 months** |  |  |  |  |  |  |
| <65 years | 1.59 (1.26–2.00)<br>(n=4338) | <0.001 | 0.94 (0.83–1.07)<br>(n=4323) | 0.374 | -0.03 (-0.04 to -0.01)<br>(n=3994) | <0.001 |
| Participants with controlled asthma† | 1.67 (0.84–3.29)<br>(n=525) | 0.142 | 0.85 (0.57–1.27)<br>(n=523) | 0.418 | -0.02 (-0.07 to 0.04)<br>(n=491) | 0.555 |
| **Association with not having received a booster vaccination in previous 6 months** |  |  |  |  |  |  |
| 65 to <75 years | 1.83 (1.51–2.23)<br>(n=2547) | <0.001 | 0.93 (0.85–1.02)<br>(n=2537) | 0.120 | -0.02 (-0.03 to -0.00)<br>(n=2399) | 0.014 |
| Clinical risk group | 1.61 (1.29–1.99)<br>(n=1885) | <0.001 | 0.96 (0.86–1.08)<br>(n=1881) | 0.513 | 0.00 (-0.02 to 0.02)<br>(n=1744) | 0.840 |
| Respiratory risk group‡ | 1.45 (1.06–1.99)<br>(n=817) | 0.021 | 0.89 (0.75–1.06)<br>(n=815) | 0.202 | -0.01 (-0.04 to 0.02)<br>(n=746) | 0.504 |

All estimates are adjusted for age and other vaccine eligibility criteria, sex, ethnicity, socioeconomic status, general health, previous SARS CoV-2 infection, and the number of vaccines received at time of infection. In all analyses, the reference group is participants who received a booster vaccine within the time frame indicated.
\*vs all other severities. †Includes all participants with asthma, who have not reported least two courses of oral corticosteroids in the preceding 24 months or at least one hospital admission for asthma in the preceding 24 months, and who are not on maintenance oral corticosteroids. Excludes participants with chronic obstructive pulmonary disease. ‡Includes participants with poorly controlled asthma or COPD. OR=odds ratio. HR=hazard ratio.

Adjusting for previous infection severity strengthened our findings on health-related quality of life but attenuated some of our findings on symptom severity, most notably when considering 6-monthly vaccination; however, small sample sizes led to reduced statistical power, particularly for the respiratory subgroups (Table S 2). Inverse probability-weighted regressions produced estimates that were largely similar to our main results, but suggested a greater effect of annual vaccination (*vs* not annual) on infection severity among participants with controlled asthma (odds ratio 2.06, 95% CI 0.97–4.36; Table S 3) and a longer duration of symptoms for participants aged 65 to <75 years when vaccinated more than 6 months prior (hazard ratio 0.91, 95% CI 0.83–1.00).

2100 participants with asthma were included in our exacerbation analysis (Figure 3), of whom 1263 (60.1%) reported at least one SARS-CoV-2 infection between October, 2021, and December, 2023. The distribution of booster vaccinations, asthma exacerbations, and infections in this cohort are shown in Figure S 1. Participants with greater average time between booster vaccinations were younger, more likely to be female, less likely to be in a clinical risk group, and reported more SARS-CoV-2 infections and asthma exacerbations during follow-up (Table S 4). Incident SARS-CoV-2 infection associated with increased risk of asthma exacerbation in boosted participants, both within a year of the last booster vaccination and later (Table 3). For severe asthma exacerbations, point estimates were higher for infections occurring more than 1 year after the last vaccination, although confidence intervals overlapped (Table 3).

**Table 3:**
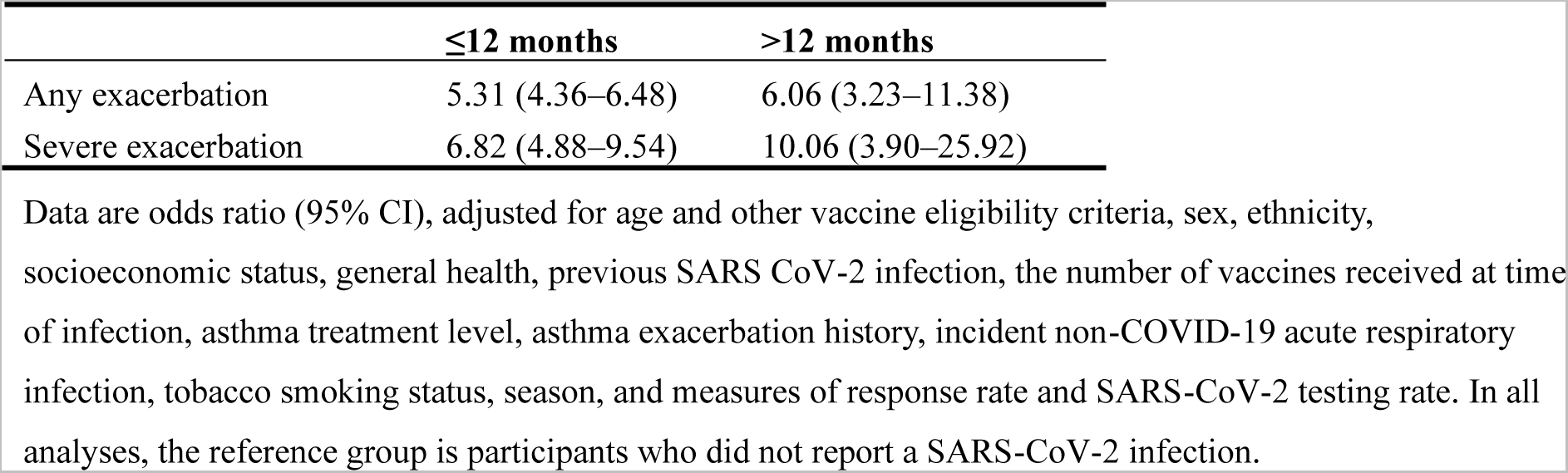
Odds of asthma exacerbations after SARS-CoV-2 infection (vs no infection) by time since vaccination.

|  | ≤12 months | >12 months |
| --- | --- | --- |
| Any exacerbation | 5.31 (4.36–6.48) | 6.06 (3.23–11.38) |
| Severe exacerbation | 6.82 (4.88–9.54) | 10.06 (3.90–25.92) |

Data are odds ratio (95% CI), adjusted for age and other vaccine eligibility criteria, sex, ethnicity, socioeconomic status, general health, previous SARS CoV-2 infection, the number of vaccines received at time of infection, asthma treatment level, asthma exacerbation history, incident non-COVID-19 acute respiratory infection, tobacco smoking status, season, and measures of response rate and SARS-CoV-2 testing rate. In all analyses, the reference group is participants who did not report a SARS-CoV-2 infection.

## Discussion

In this large, prospective, observational study of UK adults receiving booster vaccinations against SARS-CoV-2, we show that longer time since last vaccination was consistently associated with worse COVID-19 symptom severity, in all subgroups considered. Additionally, increased time since last vaccination was associated with a reduction in health-related quality of life after subsequent SARS-CoV-2 infection in participants younger than 65 years. However, we found little evidence that more recent vaccination associates with shorter symptom duration. When considering participants with asthma, incident SARS-CoV-2 infection continued to associate with asthma exacerbations in a boosted population. Importantly, our findings suggest that risk of severe exacerbations may increase with greater time since booster vaccination, although we lacked the power to show a clear effect.

Previous studies have shown decreases in vaccine effectiveness in the first 6–8 months following a booster vaccination, [17, 20, 34] accompanied by greater symptom severity when infected [14, 15] and increased risk of severe or critical COVID-19. [16-18] However, few have considered symptom duration, [14] and we are not aware of any that have looked at the effect of time since last booster vaccination on health-related quality of life. By considering these three elements of acute infection, we explore the impact of frequent booster vaccinations from multiple perspectives.

We observed a consistent relationship between self-reported symptom severity and time since vaccination; however, these results rarely translated to changes in symptom duration or health-related quality of life—and where they did, changes observed were unlikely to be clinically significant (appendix p 7). Nonetheless, the impact of reducing the severity of the acute infection—particularly among working-age adults—should not be underestimated. While 28% of our participants vaccinated within the past 6 months required bedrest, this proportion increased to 42% of those who had not been vaccinated for more than a year. Given that more than a third of UK organisations surveyed reported COVID-19 to be among the top three causes of short-term sickness absence in their workforce, [35, 36] the differences in acute COVID-19 severity we observed have the potential to translate to a huge number of working days lost.

Eligibility criteria for SARS-CoV-2 booster vaccinations in the UK have been narrowed each autumn, meaning an increasing portion of the UK population has not been vaccinated for more than a year. Among this population is people with well controlled asthma, who have not been eligible for a booster vaccine since autumn 2021. We found that incident SARS-CoV-2 infection remained associated with asthma exacerbations in boosted participants, independently of asthma severity and exacerbation history. While vaccination is known to reduce the severity of the acute infection, thus potentially reducing exacerbation risk, [37] assessment of electronic health records has shown a greater percentage of asthmatic patients being prescribed systemic corticosteroids or antibiotics when infected with Omicron as opposed to earlier variants of SARS-CoV-2, [38] suggesting that the Omicron variant may be triggering more asthma exacerbations. This is supported by paediatric studies, which have shown higher odds of asthma exacerbations with Omicron compared with other variants. [39] Therefore, despite Omicron being associated with milder disease than previous variants, [40] its impact on those at risk of asthma exacerbations remains elevated, emphasising the importance of vaccinating this population.

Although the difference was not significant, our point estimates suggest that people with asthma vaccinated more than 1 year ago may be at increased risk of severe exacerbations than those vaccinated within the past year, when infected with SARS-CoV-2. Further, well powered studies are urgently needed to assess this point conclusively, particularly in people with well-controlled asthma who have not been recently vaccinated. In particular, classification of asthma control should be made cautiously in these years following the acute pandemic, [41] as lower exposure to viruses and greater adherence to preventive behaviours means that recent exacerbation history may not be fully representative of someone’s risk of future exacerbations. [42-45] This study has several strengths. By considering symptom severity and duration, health-related quality of life, and asthma exacerbations, we present a multifaceted picture of the acute impact of breakthrough SARS-CoV-2 infection. The large size of our dataset allowed us to look at outcomes in specific subgroups of the population, according to current vaccine eligibility. Extended follow-up meant we could assess the impact of annual vaccination, whereas previous studies have mostly focused on outcomes up to 6 months post vaccination, [14-16, 18] and thus cannot accurately represent risk for populations no longer eligible for additional booster vaccinations. Additionally, our use of 6–month and 12–month thresholds, rather than shorter thresholds used in other studies, [14, 15, 34] makes it easier to interpret our findings within the context of vaccination programmes offering boosters every 6–12 months. [2] Monthly follow-up questionnaires enabled repeated measures analysis, allowing us to control for time-varying covariates when considering exacerbation risk. Detailed information on both outcomes and exposures enabled us to minimise confounding with adjusted analyses.

This study also has some limitations. With very few hospitalisations, we were unable to assess the association between time since vaccination and severe disease; however, mild or moderate COVID-19 still has a substantial impact on society, with boosted individuals taking an average of 5 days sick leave to recover. [46] We lacked power in some subgroup analyses, particularly for asthma exacerbations, meaning that further study in larger populations is still needed to confirm or refute the associations reported. We did not have data on all of the eligibility criteria for the clinical risk group, potentially leading to some misclassification of vaccine eligibility, which may have attenuated some of our results. COVIDENCE UK is a self-selected cohort, [47] meaning that some groups—such as women, older people, and those of White ethnicity—are over-represented in our study, potentially limiting the generalisability of our results. Finally, as an observational study, we cannot rule out the possibility of unmeasured or residual confounding influencing our findings. However, the richness of data in COVIDENCE UK, and use of directed acyclic graphs, means that we were able to control for a large number of potential confounders in our analyses.

In conclusion, we found that longer time since booster vaccination was consistently associated with worse symptom severity of breakthrough SARS-CoV-2 infections, but had little effect on the speed of recovery or acute changes to health-related quality of life. We also observed that incident COVID-19 continued to associate with increased risk of asthma exacerbations in a boosted population, independent of asthma severity and exacerbation history, and that risk of severe exacerbations may increase with longer time since last vaccination. These findings highlight the importance of continuing to vaccinate those who fulfil existing eligibility criteria for boosters, particularly given widespread under-vaccination, [11] and the urgent need for further research on the impacts of vaccination in people with asthma who are no longer eligible for booster vaccinations.

## Contributors

GV and ARM conceived the analysis. GV and MT contributed to data management, and have directly accessed and verified the data. Statistical analyses were done by GV, with input from PEP, ARM, MT, and SOS. GV wrote the first draft of the report. All authors provided critical conceptual input, interpreted the data analysis, and read and approved the final draft.

## Data sharing statement

De-identified participant data will be made available upon reasonable request to the corresponding author.

## Declaration of interests

PEP declares grants paid to their institution from the National Institute for Health and Care Research, UK Research and Innovation, GlaxoSmithKline, and AstraZeneca,; honoraria for lectures from AstraZeneca, GlaxoSmithKline, and Chiesi; support for attending meetings from AstraZeneca, GlaxoSmithKline, and Sanofi; advisory board participation for AstraZeneca, GlaxoSmithKline, and Sanofi; and payments to their institution for clinical trial participation by AstraZeneca, GlaxoSmithKline, Sanofi, Novartis, and Regeneron, all outside of the submitted work. The remaining authors declare no competing interests.

## Acknowledgments

COVIDENCE UK has received support from Barts Charity (MGU0459, MGU0466), Pharma Nord, the Fischer Family Foundation, DSM Nutritional Products, the Exilarch’s Foundation, the Karl R Pfleger Foundation, the AIM Foundation, Synergy Biologics, Cytoplan, the UK NIHR Clinical Research Network (52255; 52257), the Health Data Research UK BREATHE Hub, the UKRI Industrial Strategy Challenge Fund (MC_PC_19004), Thornton & Ross, Warburtons, Matthew Isaacs (personal donation), Barbara Boucher (personal donation), and Hyphens Pharma. MT was supported by Barts Charity (MGU0570). The views expressed are those of the authors and not necessarily those of the funders. We thank all participants of COVIDENCE UK, and the following organisations who supported study recruitment: Asthma UK/British Lung Foundation, the British Heart Foundation, the British Obesity Society, Cancer Research UK, Diabetes UK, Future Publishing, Kidney Care UK, Kidney Wales, Mumsnet, the National Kidney Federation, the National Rheumatoid Arthritis Society, the North West London Health Research Register (DISCOVER), Primary Immunodeficiency UK, the Race Equality Foundation, SWM Health, the Terence Higgins Trust, and Vasculitis UK.

## Supplementary material

**Table S 1:**
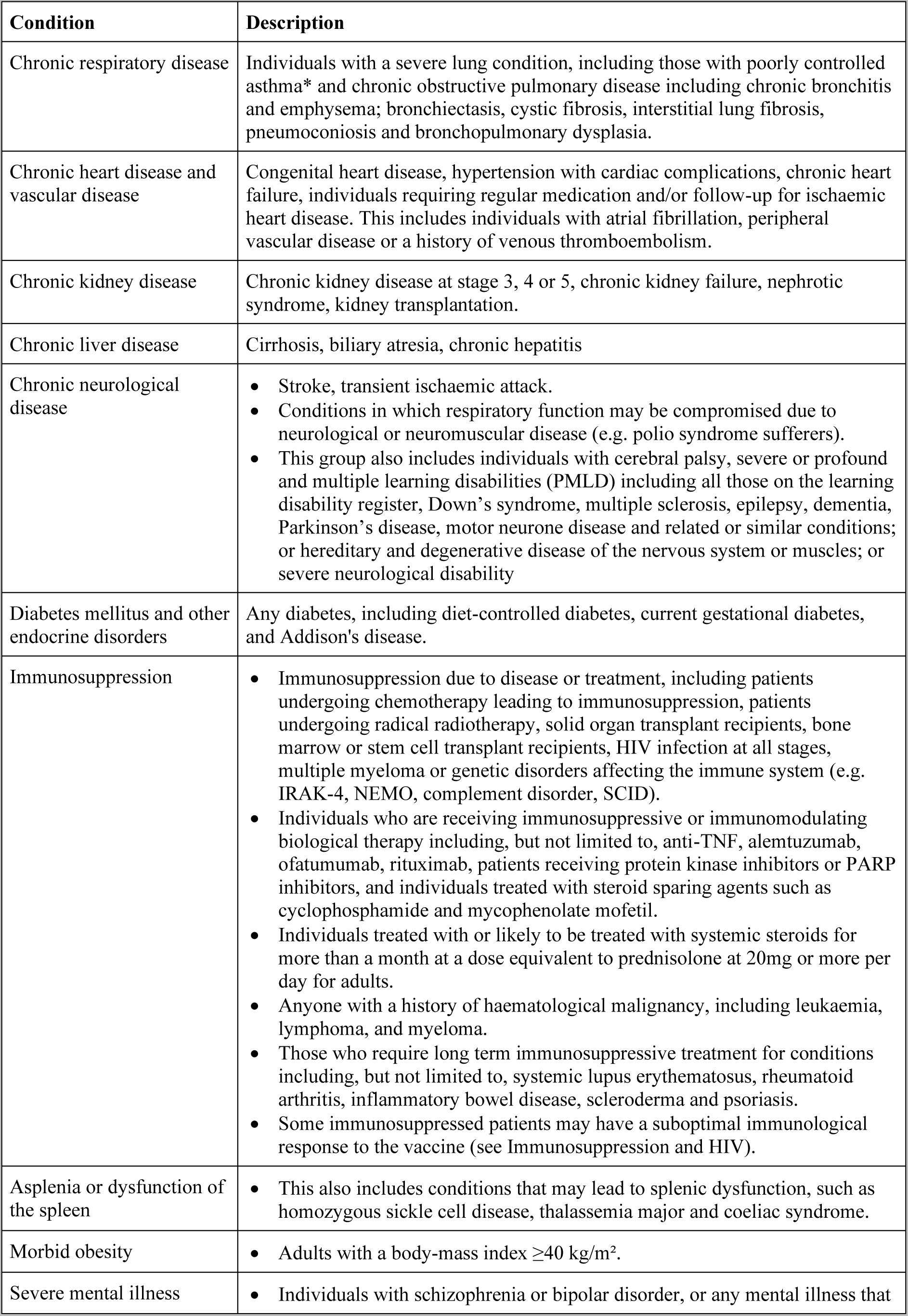
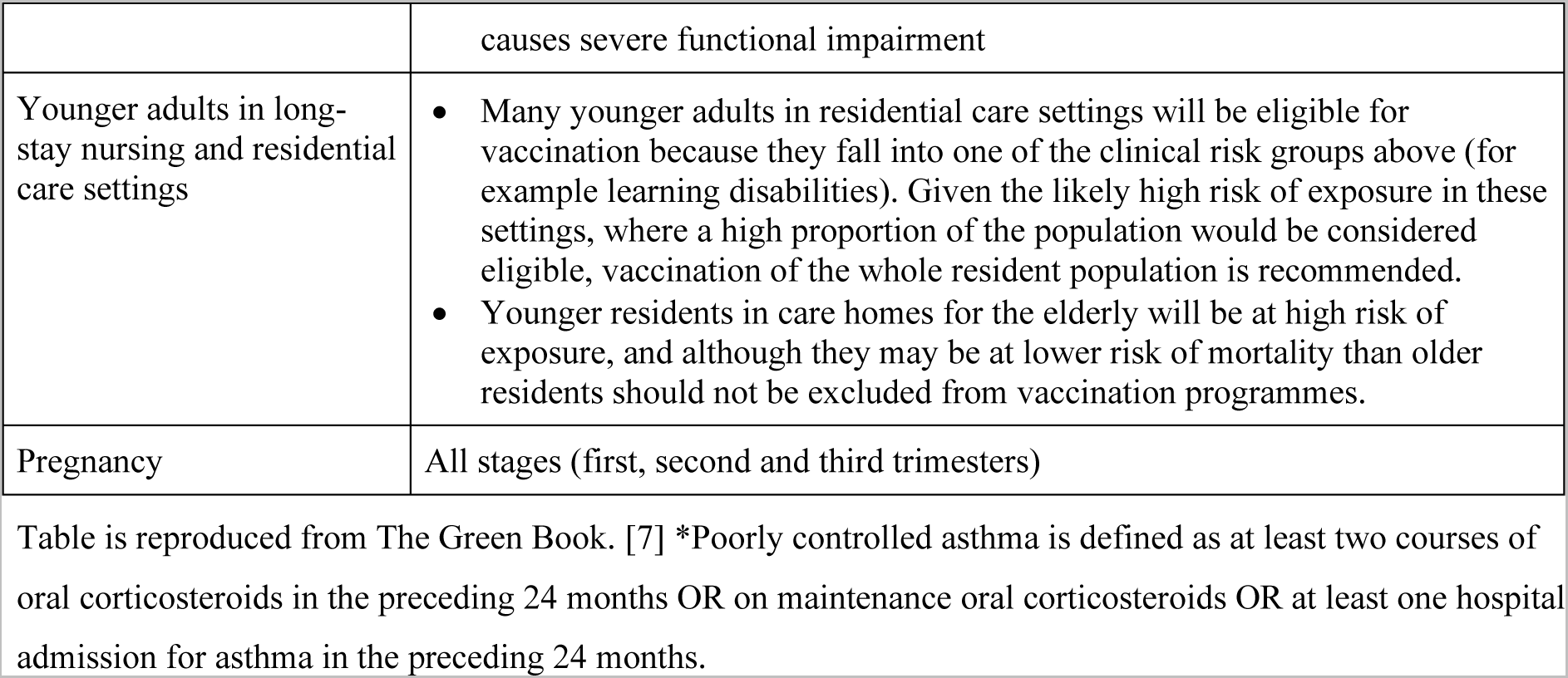
Clinical risk groups for individuals aged 16 years and over.

**Table S 2:**
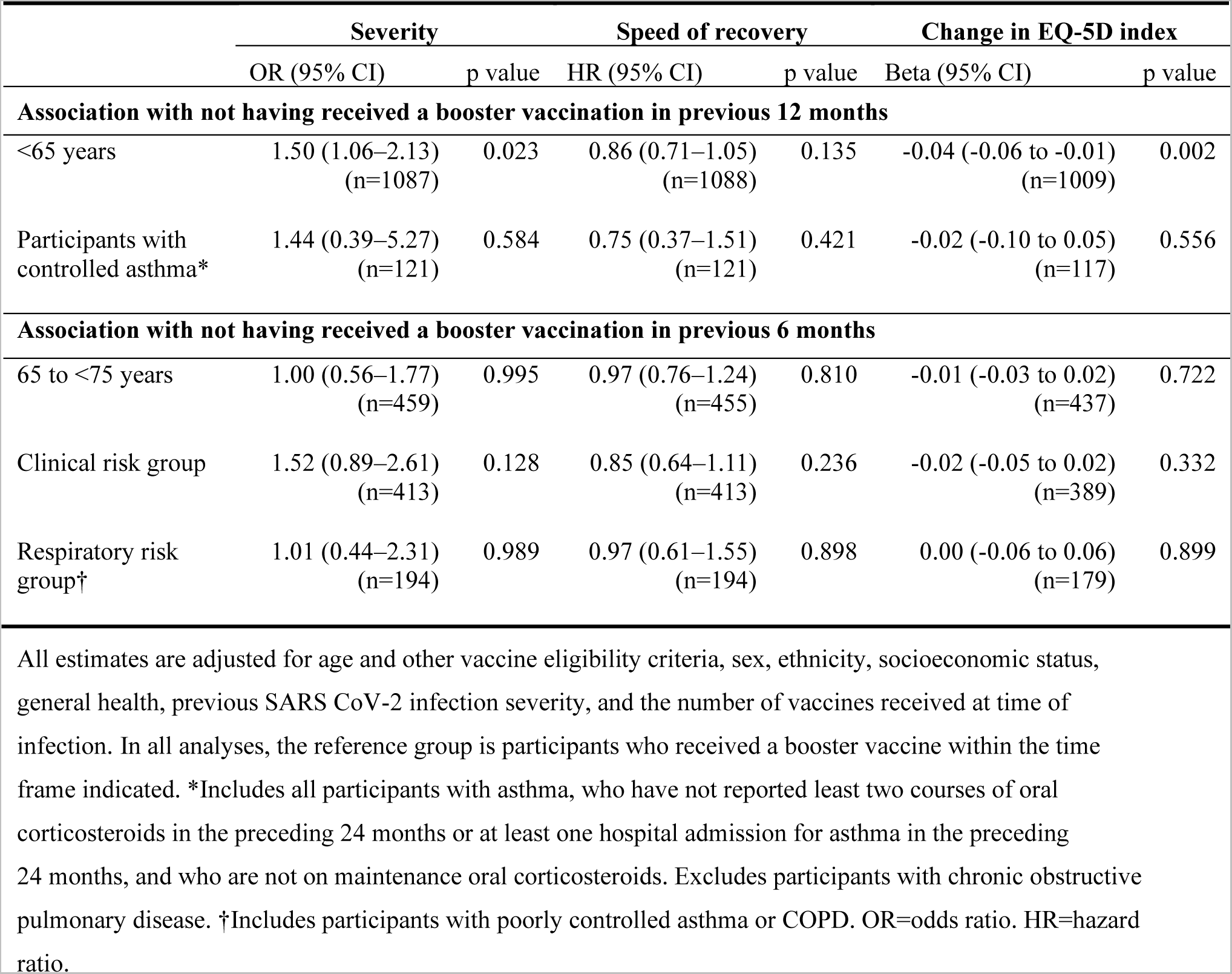
Sensitivity analysis 1: Associations between infection characteristics and time since vaccination, adjusted for previous infection severity.

**Table S 3:**
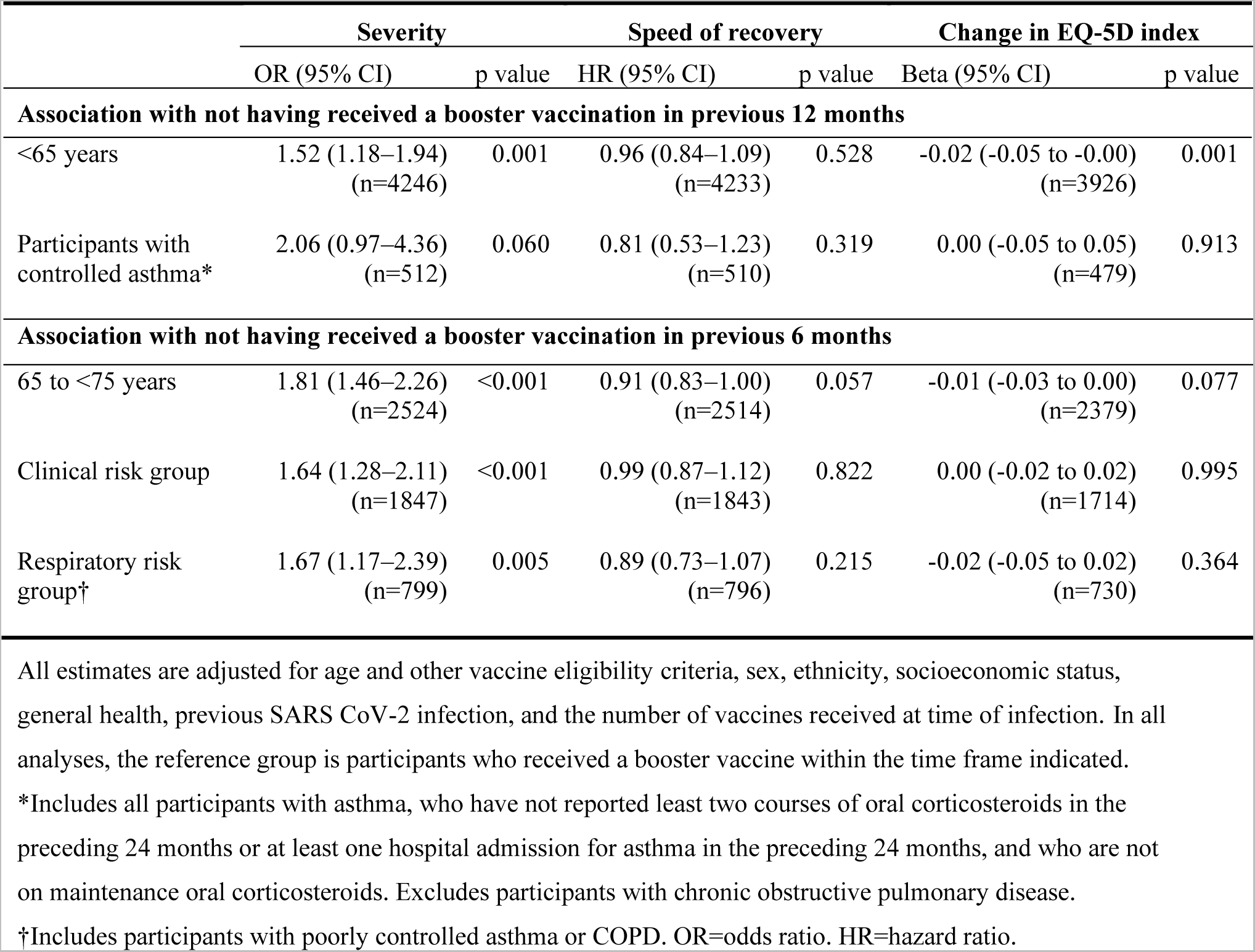
Sensitivity analysis 2: Associations between infection characteristics and time since vaccination, using inverse probability weighted regressions.

**Figure S 1:**
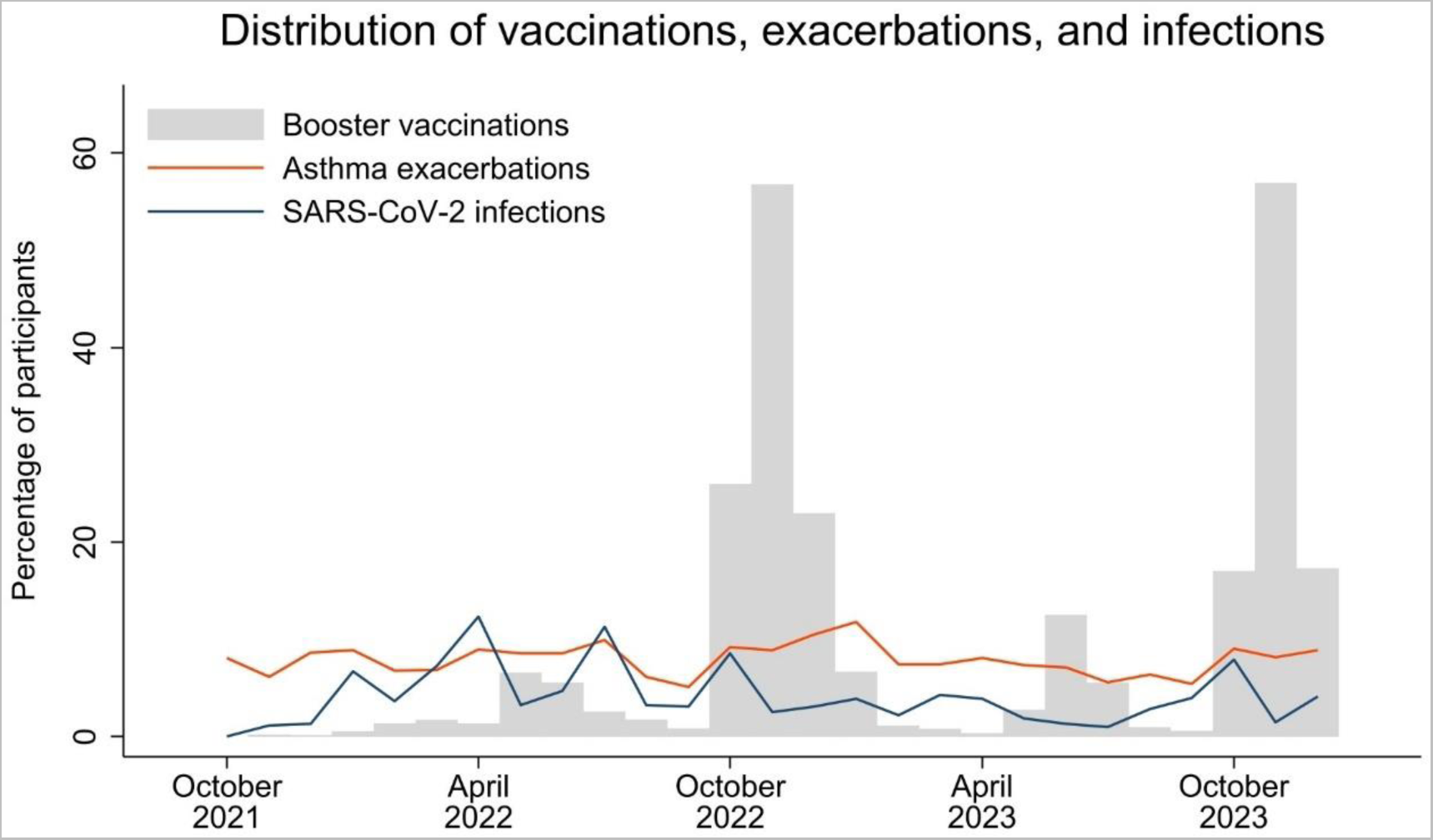
Distribution of vaccinations, exacerbations, and infections in participants with asthma

Figure shows the percentage of participants responding to each questionnaire who reported a booster vaccination, asthma exacerbation, or SARS-CoV-2 infection.

### Minimum clinically important difference

Assessment of a minimum clinically important difference (MCID) in patient-reported outcome (PRO) tools such as the EQ-5D-3L is complex, with no single established approach. [48] There are two general approaches to its determination: anchor-based and distribution-based methods. The first requires use of a so-called anchor: this can be an objective measure of improvement, but as such measures are often lacking, it is frequently a subjective measure such as a Global Assessment Rating. In order for anchor-based methods to work, the chosen anchor needs to show sufficient correlation with the PRO measure. [49] Distribution-based methods focus entirely on the distribution of the PRO measures, regardless of the patients’ perception of their progress. Various distribution-based approaches have been proposed, including the standard errors of measurement (SEM), SDs, and calculation of effect sizes. [48]

As we lack a suitable anchor to assess changes in the EQ-5D Index, we present estimates from distribution-based approaches, to help guide our interpretation of results.

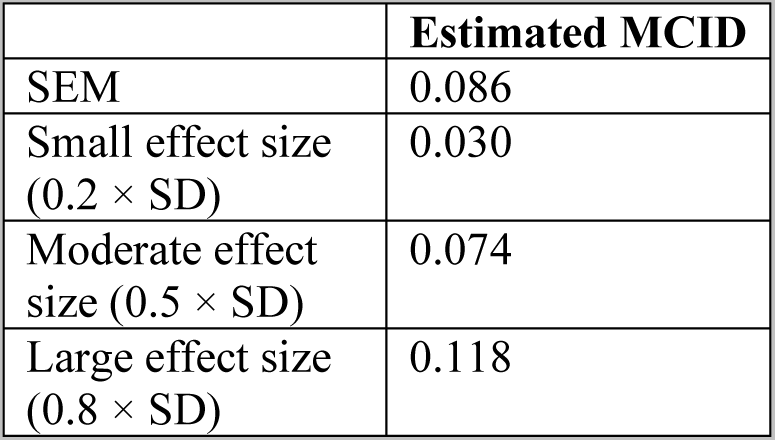

**Table S 4:**
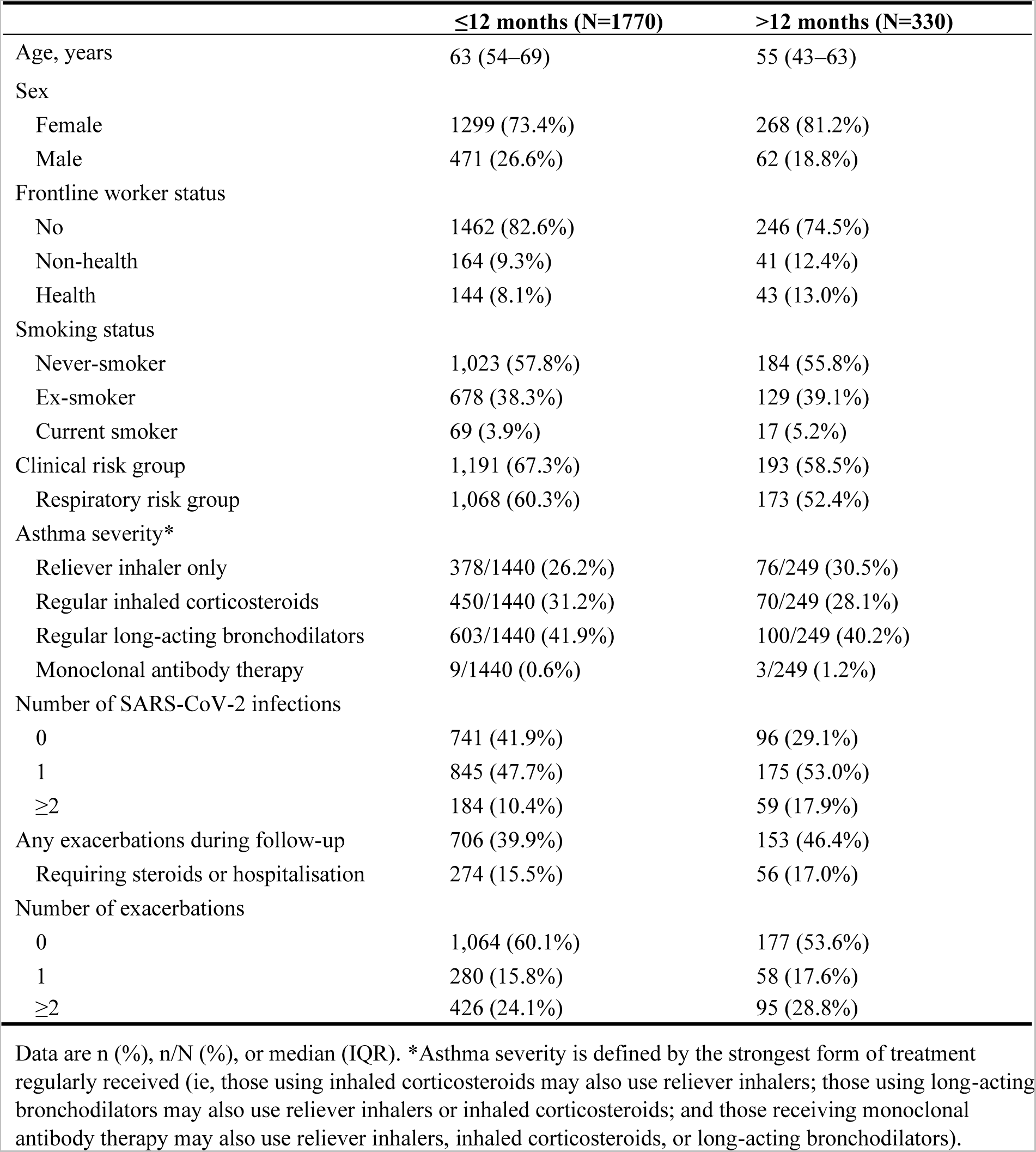
Participant characteristics for asthma analysis, by average time between booster vaccinations.

## Notes

### Author Declarations

This study is registered with ClinicalTrials.gov, NCT04330599. COVIDENCE UK was approved by Leicester South Research Ethics Committee (ref 20/EM/0117).

