## Supplementary material for "COVID-19 severity and risk of SARS-CoV-2-associated asthma exacerbation by time since booster vaccination: a longitudinal analysis of data from the COVIDENCE UK study"

*Giulia Vivaldi, Mohammad Talaei, Paul E Pfeffer, Seif O Shaheen, Adrian R Martineau*

Table S 1: Clinical risk groups for individuals aged 16 years and over

| **Condition** | **Description** |
| --- | --- |
| Chronic respiratory disease | Individuals with a severe lung condition, including those with poorly controlled asthma* and chronic obstructive pulmonary disease including chronic bronchitis and emphysema; bronchiectasis, cystic fibrosis, interstitial lung fibrosis, pneumoconiosis and bronchopulmonary dysplasia. |
| Chronic heart disease and vascular disease | Congenital heart disease, hypertension with cardiac complications, chronic heart failure, individuals requiring regular medication and/or follow-up for ischaemic heart disease. This includes individuals with atrial fibrillation, peripheral vascular disease or a history of venous thromboembolism. |
| Chronic kidney disease | Chronic kidney disease at stage 3, 4 or 5, chronic kidney failure, nephrotic syndrome, kidney transplantation. |
| Chronic liver disease | Cirrhosis, biliary atresia, chronic hepatitis |
| Chronic neurological disease | - Stroke, transient ischaemic attack. - Conditions in which respiratory function may be compromised due to neurological or neuromuscular disease (e.g. polio syndrome sufferers). - This group also includes individuals with cerebral palsy, severe or profound and multiple learning disabilities (PMLD) including all those on the learning disability register, Down’s syndrome, multiple sclerosis, epilepsy, dementia, Parkinson’s disease, motor neurone disease and related or similar conditions; or hereditary and degenerative disease of the nervous system or muscles; or severe neurological disability |
| Diabetes mellitus and other endocrine disorders | Any diabetes, including diet-controlled diabetes, current gestational diabetes, and Addison's disease. |
| Immunosuppression | - Immunosuppression due to disease or treatment, including patients undergoing chemotherapy leading to immunosuppression, patients undergoing radical radiotherapy, solid organ transplant recipients, bone marrow or stem cell transplant recipients, HIV infection at all stages, multiple myeloma or genetic disorders affecting the immune system (e.g. IRAK-4, NEMO, complement disorder, SCID). - Individuals who are receiving immunosuppressive or immunomodulating biological therapy including, but not limited to, anti-TNF, alemtuzumab, ofatumumab, rituximab, patients receiving protein kinase inhibitors or PARP inhibitors, and individuals treated with steroid sparing agents such as cyclophosphamide and mycophenolate mofetil. - Individuals treated with or likely to be treated with systemic steroids for more than a month at a dose equivalent to prednisolone at 20mg or more per day for adults. - Anyone with a history of haematological malignancy, including leukaemia, lymphoma, and myeloma. - Those who require long term immunosuppressive treatment for conditions including, but not limited to, systemic lupus erythematosus, rheumatoid arthritis, inflammatory bowel disease, scleroderma and psoriasis. - Some immunosuppressed patients may have a suboptimal immunological response to the vaccine (see Immunosuppression and HIV). |
| Asplenia or dysfunction of the spleen | - This also includes conditions that may lead to splenic dysfunction, such as homozygous sickle cell disease, thalassemia major and coeliac syndrome. |
| Morbid obesity | - Adults with a body-mass index ≥40 kg/m². |
| Severe mental illness | - Individuals with schizophrenia or bipolar disorder, or any mental illness that causes severe functional impairment |
| Younger adults in long-stay nursing and residential care settings | - Many younger adults in residential care settings will be eligible for vaccination because they fall into one of the clinical risk groups above (for example learning disabilities). Given the likely high risk of exposure in these settings, where a high proportion of the population would be considered eligible, vaccination of the whole resident population is recommended. - Younger residents in care homes for the elderly will be at high risk of exposure, and although they may be at lower risk of mortality than older residents should not be excluded from vaccination programmes. |
| Pregnancy | All stages (first, second and third trimesters) |

Table is reproduced from The Green Book. [7] *Poorly controlled asthma is defined as at least two courses of oral corticosteroids in the preceding 24 months OR on maintenance oral corticosteroids OR at least one hospital admission for asthma in the preceding 24 months.

Table S 2: Sensitivity analysis 1: Associations between infection characteristics and time since vaccination, adjusted for previous infection severity

|  | **Severity** | | **Speed of recovery** | | **Change in EQ-5D index** | |
| --- | --- | --- | --- | --- | --- | --- |
|  | OR (95% CI) | p value | HR (95% CI) | p value | Beta (95% CI) | p value |
| **Association with not having received a booster vaccination in previous 12 months** | | | | | | |
| <65 years | 1.50 (1.06–2.13) (n=1087) | 0.023 | 0.86 (0.71–1.05) (n=1088) | 0.135 | -0.04 (-0.06 to -0.01) (n=1009) | 0.002 |
| Participants with controlled asthma* | 1.44 (0.39–5.27) (n=121) | 0.584 | 0.75 (0.37–1.51) (n=121) | 0.421 | -0.02 (-0.10 to 0.05) (n=117) | 0.556 |
| **Association with not having received a booster vaccination in previous 6 months** | | | | | | |
| 65 to <75 years | 1.00 (0.56–1.77) (n=459) | 0.995 | 0.97 (0.76–1.24) (n=455) | 0.810 | -0.01 (-0.03 to 0.02) (n=437) | 0.722 |
| Clinical risk group | 1.52 (0.89–2.61) (n=413) | 0.128 | 0.85 (0.64–1.11) (n=413) | 0.236 | -0.02 (-0.05 to 0.02) (n=389) | 0.332 |
| Respiratory risk group† | 1.01 (0.44–2.31) (n=194) | 0.989 | 0.97 (0.61–1.55) (n=194) | 0.898 | 0.00 (-0.06 to 0.06) (n=179) | 0.899 |

All estimates are adjusted for age and other vaccine eligibility criteria, sex, ethnicity, socioeconomic status, general health, previous SARS CoV-2 infection severity, and the number of vaccines received at time of infection. In all analyses, the reference group is participants who received a booster vaccine within the time frame indicated. *Includes all participants with asthma, who have not reported least two courses of oral corticosteroids in the preceding 24 months or at least one hospital admission for asthma in the preceding 24 months, and who are not on maintenance oral corticosteroids. Excludes participants with chronic obstructive pulmonary disease. †Includes participants with poorly controlled asthma or COPD. OR=odds ratio. HR=hazard ratio.

Table S 3: Sensitivity analysis 2: Associations between infection characteristics and time since vaccination, using inverse probability weighted regressions

|  | **Severity** | | **Speed of recovery** | | **Change in EQ-5D index** | |
| --- | --- | --- | --- | --- | --- | --- |
|  | OR (95% CI) | p value | HR (95% CI) | p value | Beta (95% CI) | p value |
| **Association with not having received a booster vaccination in previous 12 months** | | | | | | |
| <65 years | 1.52 (1.18–1.94) (n=4246) | 0.001 | 0.96 (0.84–1.09) (n=4233) | 0.528 | -0.02 (-0.05 to -0.00) (n=3926) | 0.001 |
| Participants with controlled asthma* | 2.06 (0.97–4.36) (n=512) | 0.060 | 0.81 (0.53–1.23) (n=510) | 0.319 | 0.00 (-0.05 to 0.05) (n=479) | 0.913 |
| **Association with not having received a booster vaccination in previous 6 months** | | | | | | |
| 65 to <75 years | 1.81 (1.46–2.26) (n=2524) | <0.001 | 0.91 (0.83–1.00) (n=2514) | 0.057 | -0.01 (-0.03 to 0.00) (n=2379) | 0.077 |
| Clinical risk group | 1.64 (1.28–2.11) (n=1847) | <0.001 | 0.99 (0.87–1.12) (n=1843) | 0.822 | 0.00 (-0.02 to 0.02) (n=1714) | 0.995 |
| Respiratory risk group† | 1.67 (1.17–2.39) (n=799) | 0.005 | 0.89 (0.73–1.07) (n=796) | 0.215 | -0.02 (-0.05 to 0.02) (n=730) | 0.364 |

All estimates are adjusted for age and other vaccine eligibility criteria, sex, ethnicity, socioeconomic status, general health, previous SARS CoV-2 infection, and the number of vaccines received at time of infection. In all analyses, the reference group is participants who received a booster vaccine within the time frame indicated. *Includes all participants with asthma, who have not reported least two courses of oral corticosteroids in the preceding 24 months or at least one hospital admission for asthma in the preceding 24 months, and who are not on maintenance oral corticosteroids. Excludes participants with chronic obstructive pulmonary disease. †Includes participants with poorly controlled asthma or COPD. OR=odds ratio. HR=hazard ratio.

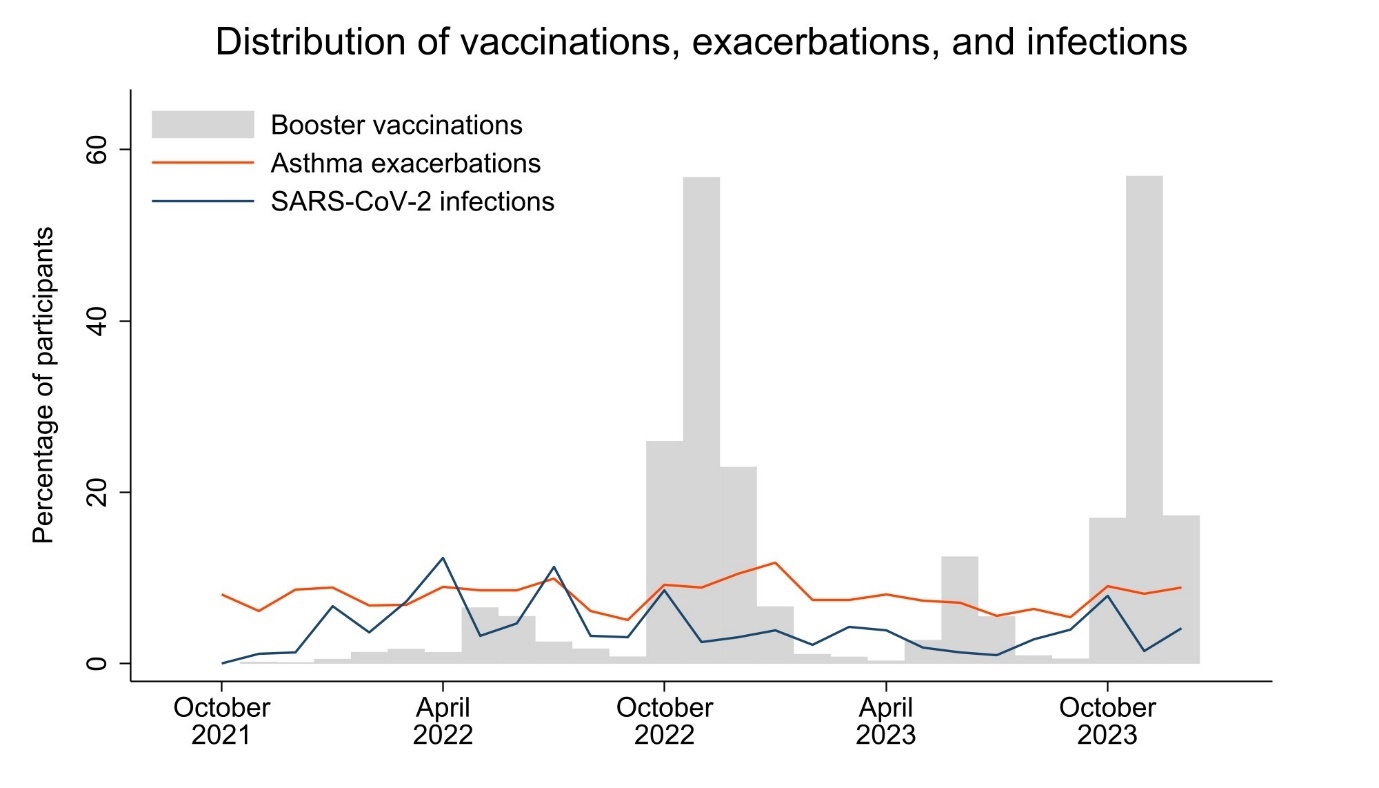

Figure S 1: Distribution of vaccinations, exacerbations, and infections in participants with asthma

Figure shows the percentage of participants responding to each questionnaire who reported a booster vaccination, asthma exacerbation, or SARS-CoV-2 infection.

Minimum clinically important difference

Assessment of a minimum clinically important difference (MCID) in patient-reported outcome (PRO) tools such as the EQ-5D-3L is complex, with no single established approach. [48] There are two general approaches to its determination: anchor-based and distribution-based methods. The first requires use of a so-called anchor: this can be an objective measure of improvement, but as such measures are often lacking, it is frequently a subjective measure such as a Global Assessment Rating. In order for anchor-based methods to work, the chosen anchor needs to show sufficient correlation with the PRO measure. [49] Distribution-based methods focus entirely on the distribution of the PRO measures, regardless of the patients’ perception of their progress. Various distribution-based approaches have been proposed, including the standard errors of measurement (SEM), SDs, and calculation of effect sizes. [48]

As we lack a suitable anchor to assess changes in the EQ-5D Index, we present estimates from distribution-based approaches, to help guide our interpretation of results.

|  | **Estimated MCID** |
| --- | --- |
| SEM | 0.086 |
| Small effect size (0.2 × SD) | 0.030 |
| Moderate effect size (0.5 × SD) | 0.074 |
| Large effect size (0.8 × SD) | 0.118 |

Table S 4: Participant characteristics for asthma analysis, by average time between booster vaccinations

|  | **≤12 months (N=1770)** | **>12 months (N=330)** |
| --- | --- | --- |
| Age, years | 63 (54–69) | 55 (43–63) |
| Sex |  |  |
| Female | 1299 (73.4%) | 268 (81.2%) |
| Male | 471 (26.6%) | 62 (18.8%) |
| Frontline worker status |  |  |
| No | 1462 (82.6%) | 246 (74.5%) |
| Non-health | 164 (9.3%) | 41 (12.4%) |
| Health | 144 (8.1%) | 43 (13.0%) |
| Smoking status |  |  |
| Never-smoker | 1,023 (57.8%) | 184 (55.8%) |
| Ex-smoker | 678 (38.3%) | 129 (39.1%) |
| Current smoker | 69 (3.9%) | 17 (5.2%) |
| Clinical risk group | 1,191 (67.3%) | 193 (58.5%) |
| Respiratory risk group | 1,068 (60.3%) | 173 (52.4%) |
| Asthma severity* |  |  |
| Reliever inhaler only | 378/1440 (26.2%) | 76/249 (30.5%) |
| Regular inhaled corticosteroids | 450/1440 (31.2%) | 70/249 (28.1%) |
| Regular long-acting bronchodilators | 603/1440 (41.9%) | 100/249 (40.2%) |
| Monoclonal antibody therapy | 9/1440 (0.6%) | 3/249 (1.2%) |
| Number of SARS-CoV-2 infections |  |  |
| 0 | 741 (41.9%) | 96 (29.1%) |
| 1 | 845 (47.7%) | 175 (53.0%) |
| ≥2 | 184 (10.4%) | 59 (17.9%) |
| Any exacerbations during follow-up | 706 (39.9%) | 153 (46.4%) |
| Requiring steroids or hospitalisation | 274 (15.5%) | 56 (17.0%) |
| Number of exacerbations |  |  |
| 0 | 1,064 (60.1%) | 177 (53.6%) |
| 1 | 280 (15.8%) | 58 (17.6%) |
| ≥2 | 426 (24.1%) | 95 (28.8%) |

Data are n (%), n/N (%), or median (IQR). *Asthma severity is defined by the strongest form of treatment regularly received (ie, those using inhaled corticosteroids may also use reliever inhalers; those using long-acting bronchodilators may also use reliever inhalers or inhaled corticosteroids; and those receiving monoclonal antibody therapy may also use reliever inhalers, inhaled corticosteroids, or long-acting bronchodilators).
